# Anemia during pregnancy and adverse maternal outcomes in Georgia – a birth registry-based cohort study

**DOI:** 10.1101/2023.11.10.23298382

**Authors:** Natia Skhvitaridze, Amiran Gamkrelidze, Tinatin Manjavidze, Tormod Brenn, Erik Anda, Charlotta Rylander

## Abstract

**Background:** Anemia in pregnancy is an important public health challenge; however, it has not been thoroughly studied in Georgia. We assessed the prevalence of anemia during pregnancy across Georgia and the association between anemia in the third trimester of pregnancy and adverse maternal outcomes.

**Methods:** We used data from the Georgian Birth Registry and included pregnant women who delivered between January 1, 2019, and August 31, 2022 (n=158,668). The prevalence of anemia (hemoglobin (Hb) < 110 g/L) at any time during pregnancy was calculated per region. Women in the third trimester were classified into three groups, based on their lowest measured Hb value: no (Hb ≥110 g/L, reference group); mild (Hb 100-109 g/L); and moderate to severe anemia (Hb <99 g/L). Adjusted odds ratios (aOR) with 95% confidence intervals (CIs) were calculated for the associations between anemia status and post-delivery intensive care unit (ICU) admission and preterm delivery.

**Results:** The prevalence of anemia occurring at least once during pregnancy was 40.6%, with large regional differences in anemia prevalence (25.1%–47.0%). Of 105,811 pregnant women with Hb measurements in the third trimester, 71.0% had no anemia; 20.9%, mild anemia; and 8.1%, moderate or severe anemia. The odds of post-delivery ICU admission did not increase linearly with decreasing Hb value (*P* for trend .13), and the relationship was inverse for preterm delivery (*P* for trend .01).

**Conclusions:** A considerable proportion of pregnant women in Georgia have anemia during pregnancy, and the prevalence and quality of reporting differ across regions. Anemia occurring in the third trimester did not substantially increase the odds of maternal ICU admission or preterm delivery. To accelerate national progress toward the Sustainable Development Goals and mitigate the consequences of anemia, equal countrywide access to high-quality antenatal care programs and complete registration of Hb values should be ensured.

## Introduction

Anemia in pregnancy, defined as a hemoglobin (Hb) level below 110 g/L [1], is an important public health challenge, affecting 30-60% of pregnancies globally, and mainly women in the third trimester [2–4]. Iron deficiency, generally due to a low dietary iron intake, is the most common cause of anemia. It has been estimated that iron deficiency anemia contributes almost 22% to maternal deaths worldwide, most occurring in low-income countries [5]. Several systematic reviews and meta-analyses have reported that anemia in pregnancy significantly increases the risk of preterm delivery [6–8]. Moreover, increased risk of maternal mortality and other adverse outcomes, such as severe post-partum hemorrhage, maternal shock, and admission to the post-delivery intensive care unit (ICU), have also been associated with anemia [9–11]. However, recent studies have reported both positive and inverse associations between anemia in pregnancy and several adverse maternal outcomes. This could be due to differences in screening, follow-up approaches, and treatment regimens, and the contradictory results pose significant challenges for decision- and policymakers in implementing evidence-based recommendations [2, 12, 13].

The diagnosis of anemia is straightforward but the disease is silent, with few physical symptoms, and it is much more complex to identify the underlying causes of anemia [14]. Many women in low- and middle-income countries may also have had undiagnosed anemia before the pregnancy [13]. Initiated by the World Health Organization (WHO), the World Health Assembly approved global targets for maternal, infant, and young child nutrition in 2012 with a commitment to reduce the prevalence of anemia in women of reproductive age by 50% before 2025 [15]. In 2020, ending anemia in women aged 15–49 was added to the Sustainable Development Goals indicator 2.2.3 [16]. The deadline for the global targets has been extended to 2030 [17]. Despite this, knowledge about global progress toward the anemia target is scarce, and global comparisons are challenged by differences in the reporting of anemia prevalence across countries [18]. Thus, the disease remains underdiagnosed and understudied [19].

Georgia is an upper-middle-income country in the Caucasus with a population of 3.7 million [20]. Reducing maternal anemia and improving reproductive healthcare are national priorities in Georgia. However, little is known about the prevalence of anemia in the country and its impact on maternal morbidity. The most recent study on this subject is the Georgian National Nutrition Survey conducted in 2009, in which 25.6% of pregnant women had anemia [21]. This study aimed to provide information about the national and regional prevalence of anemia in pregnant women in Georgia and to evaluate the associations between anemia in the third trimester and maternal transfer to the post-delivery ICU and preterm delivery.

## Materials and methods

### The Georgian Birth Registry

Following World Health Organization recommendations, eight antenatal care (ANC) visits are free of charge for Georgian citizens [22]. Most pregnant women in Georgia attend at least one ANC visit (95.8% in 2021) and deliver in a health facility (99.8% in 2021) [23]. The Georgian Birth Registry (GBR) contains countrywide information of all medical facility-based deliveries and ANC visits [24, 25]. The coverage of newborns registered in the GBR was 99.8% in 2021 [26]. Registering information in the GBR is mandatory for the involved healthcare facilities. The GBR includes many variables, including demographic characteristics, disease history, and information about the current pregnancy up to post-delivery hospital discharge [26]. The GBR also includes Hb levels registered during ANC visits. According to national guidelines, the first ANC visit, recommended before gestational age (GA) week 13, should include a full blood count, including Hb measurement. Hb levels should be measured again at the third ANC visit at GA week 26, the fourth ANC visit at GA week 30, and the sixth ANC visit at GA week 36 [27]. The study data were accessed for research purposes on July 12, 2023.

### Study sample

We initially included the data of all women registered in the GBR who gave birth between January 1, 2019, and August 31, 2022, who were at least at GA 22 weeks (n=166,043). We excluded women who did not attend ANC during their pregnancy (n=7,375), resulting in an analytical sample of 158, 668 women. Of these, 28,709 women attended ANC, but had no or no reliable Hb measure registered in the GBR. To calculate the national and regional prevalence of anemia among women who delivered after week 22, we used the data of all women who had at least one Hb measurement during the pregnancy (n=129,959). To study the association between anemia severity, post-delivery ICU admission, and preterm delivery, we included only women with at least one Hb measurement in the third trimester (GA week 28 and onwards; n=105,811 women). For the third trimester cutoff, we followed the recommendations of the National Institute for Health and Care Excellence, UK [28] (Fig. 1).

**Figure 1.**
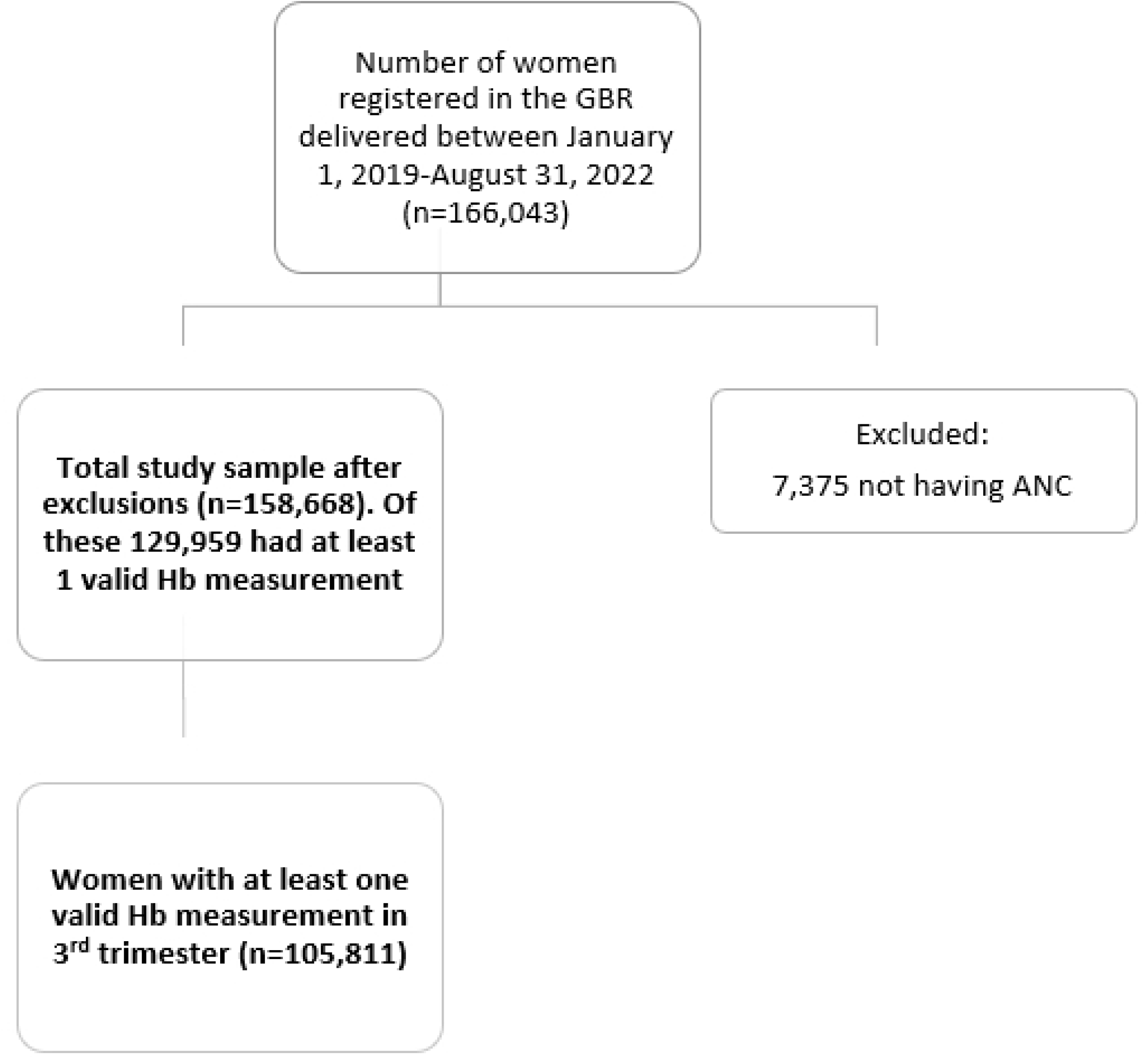
Flow chart of the study sample.

### Exposure and covariates

Hb levels ≤40 g/L or ≥180 g/L were considered implausible [29] and were recoded as “missing.” Women were considered to have anemia at any time during pregnancy if their lowest recorded Hb level was <110 g/L. We classified pregnant women into anemia severity groups based on their lowest measured Hb value at any time during pregnancy (total study sample) or during the third trimester (sub-sample used for the regression analysis): no anemia (Hb ≥110 g/L, reference group); mild anemia (Hb 100–109 g/L); moderate (Hb 70–99 g/L) and severe (Hb <70 g/L) anemia. The moderate and severe anemia groups were combined in the regression analysis because of the limited number of women in the severe anemia group. The thresholds for Hb levels were based on the WHO recommendation for anemia classification [1].

Information on sociodemographic characteristics (maternal age, residency, and education), year of delivery, obstetric history (frequency of ANC, parity, plurality, mode of delivery, and bleeding during pregnancy), body mass index (BMI) at the first ANC visit before GA week 13, and GA at the lowest recorded Hb value were extracted from the GBR. Maternal age was defined as the mother’s age at delivery. Residence was categorized as rural, urban, or unknown, and education was classified as primary, secondary, higher, or unknown. We dichotomized most covariates related to obstetric history: parity as primiparous or multiparous, plurality as singleton or multiple, and bleeding during pregnancy as yes or no, based on the International Classification of Disease version 10 (ICD10) code O 20.0 [30]. Number of ANC visits was divided into three groups (<4 visits; 4-8 visits; >8 visits), and the mode of delivery was grouped into two - cesarean section or vaginal delivery. BMI at the first ANC visit <GA week 13 was divided into five groups according to the WHO classification of body weight status (<18.5 kg/m^2^, 18.5–24.9 kg/m^2^, 25–30 kg/m^2^, >30 kg/m^2^ and missing BMI before GA week 13) [31].

### Outcomes

We extracted information on the two main outcomes, post-delivery ICU admission and preterm delivery from the GBR. Post-delivery ICU admission was defined as any admission to the post-delivery ICU during or after delivery. Preterm delivery was defined as delivery before GA week 37.

### Ethics and consent

Regional Committees for Medical and Health Research Ethics in Northern Norway approved the study protocol (Ref: 577179, 20/02/2023). All data included in this study were extracted from the GBR and anonymized before the researchers received the data. Registration in the national registries is mandatory by law, and citizens cannot refuse registration. Consent was not obtained from the study participants as it is deemed unnecessary according to national regulations [32]. The study reporting is in line with the Helsinki declaration and the strengthening the reporting of observational studies in epidemiology (STROBE) guideline.

### Statistical analysis

We calculated the period prevalence (2019–2022) of anemia at any time during pregnancy per region by dividing the number of pregnant women in each region with at least one Hb measurement of < 110 g/L (anemia cases) by the total number of women in that region with at least one valid Hb measurement registered in the GBR during the same study period. Descriptive statistics for anemia testing status are presented as frequencies and percentages for nominal variables and means and standard deviations for continuous variables.

Binary logistic regression assessed the associations between anemia (three exposure groups) in the third trimester and post-delivery ICU admission and preterm delivery. We estimated crude associations and associations adjusted for confounding factors. We drew directed acyclic graphs (DAGs) to identify possible confounding factors between anemia during pregnancy and post-delivery ICU admission and preterm delivery. The relationships depicted between the variables included in the DAGs are based on previous literature and the underlying theory (details are described in Supplementary file 1 and 2). The DAGs assumed that anemia had a causal effect on the outcomes. Based on the DAGs, the post-delivery ICU admission model was adjusted for age, BMI, bleeding during pregnancy, cesarean section delivery, and parity, and the preterm delivery model was adjusted for age, education, BMI, bleeding during pregnancy, and plurality. The results were presented as crude and adjusted odds ratios (aORs) with 95% confidence intervals (CIs). We included continuous Hb values in the logistic regression models to test for linear trends and extracted the *P* value, corresponding to one incremental change in Hb. The significance threshold was set at .05. All statistical analyses were performed using STATA version 17 (StataCorp, TX, USA).

## Results

During the study period, 52,826 pregnant women had anemia (Hb <110 g/L) at least once during pregnancy, translating into a prevalence of 40.6%. Of these, 28.3%, 11.9 %, and 0.5% had mild, moderate, and severe anemia, respectively (Supplementary Table 1). Only 10,884 (8.4%) pregnant women had an Hb measurement before GA week 13, and 28,709 (18.1%) pregnant women had no Hb measurement during the entire pregnancy, although they attended ANC (Supplementary Table 1). Having two Hb measurements during pregnancy was the most common scenario (n=67,425, 51.9%).

The prevalence of anemia differed across the 12 administrative regions in Georgia and was lowest in Samtskhe-Javakheti (25.1%) and Racha (34.4%) and highest in Adjara (47.0%) and Kvemo Kartli (46.5%) (Fig. 2).

**Figure 2.**
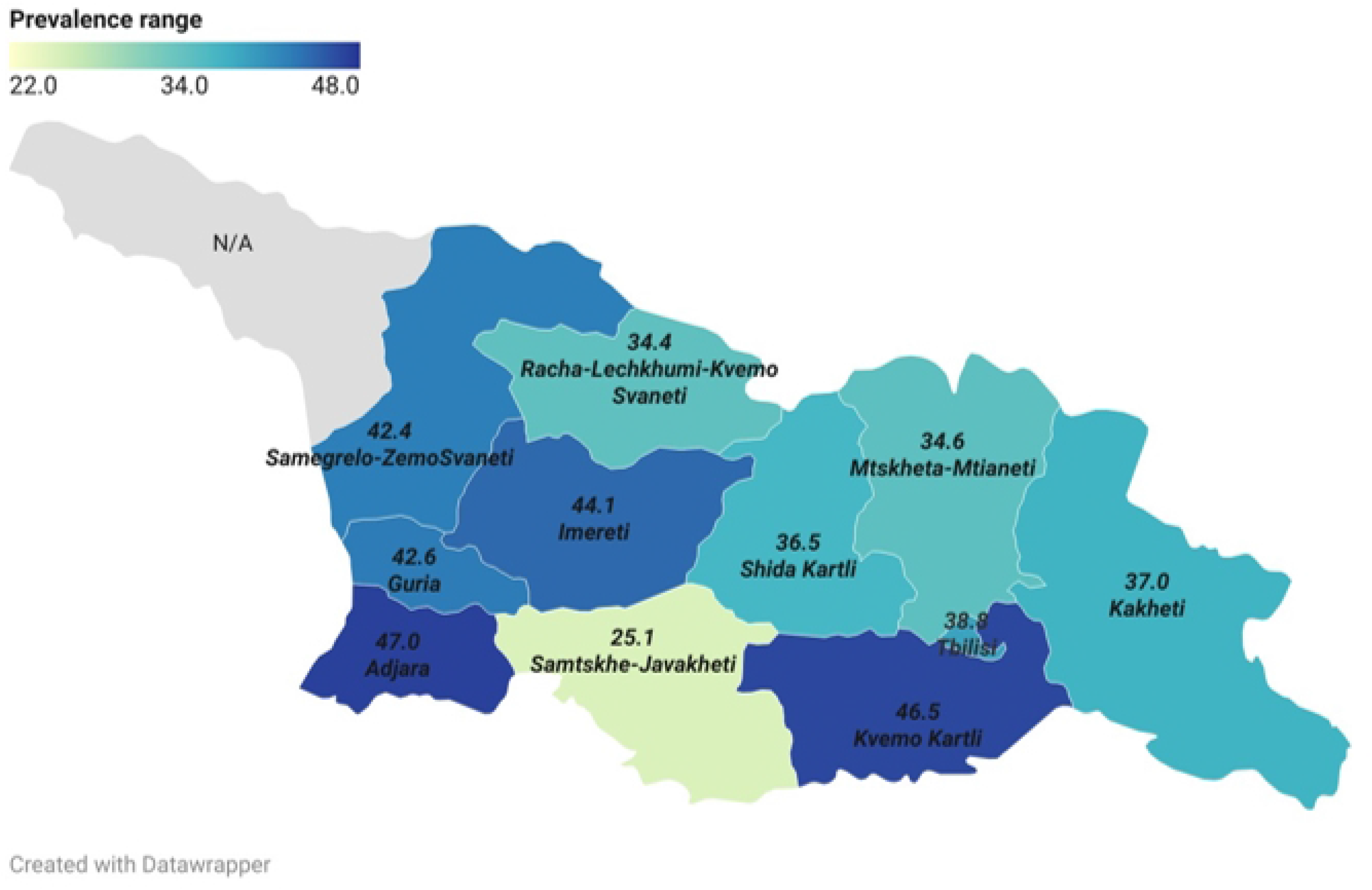
Anemia prevalence in Georgian regions, 2019-2022.

The proportion of women with no or no reliable Hb measurements during pregnancy also varied considerably across regions; it was highest in Samegrelo and Zemo Svaneti, where 33.3% of pregnant women had no registered Hb measurement and lowest in Samtskhe-Javakheti (7.1%) and Imereti (8.0%) (Supplementary Table 2). The number of women who did not attend ANC also varied by region; Mtskheta-Mtianeti and Kakheti had the highest proportions of non-attendance (6.6% and 6.1%, respectively) and Adjara, Imereti, and Samtskhe-Javakheti had the lowest proportions (2.6%) (Supplementary Table 3).

Of the 105,811 included women with Hb measurement in their third trimester, 71.0% had no anemia, 20.9% had mild anemia, and 8.1% had moderate or severe anemia (Table 1). The third trimester prevalence of anemia varied by year and was highest in 2019 (25.9% and 11.5% for mild and moderate/severe anemia, respectively) and lowest in 2022 (17.3% and 5.9% for mild and moderate/severe anemia, respectively). The mean maternal age was around 28 years for all anemia groups. Most women lived in urban areas, had secondary education, attended between four and eight ANC visits, were normal weight, multipara, had a singleton pregnancy, and delivered vaginally. Notably, mild anemia was mostly diagnosed at GA week 35, and moderate/severe at GA week 34. Women with anemia in the third trimester were more likely to have more than eight ANC visits, have a BMI below 18.5 kg/m^2^ at GA week <13, and be multiparous compared to women with no anemia in the third trimester. Bleeding during pregnancy was rare in all pregnant women and was almost equally distributed across all three anemia groups. Characteristics according to anemia status for the total study sample (Supplementary Table 1) were similar to those of the subsample of women with Hb measurement in the third trimester.

**Table 1.** Maternal Baseline Characteristics by Anemia Status During Third Trimester of Pregnancy.

| Characteristics | No anemia<br>>110 g/L | Mild<br>100-109 g/L | Moderate & Severe<br><99 g/L |
| --- | --- | --- | --- |
| n (row, %) | 75,114 (71.0) | 22,122 (20.9) | 8,575 (8.1) |
| Year of delivery, n (row, %) |  |  |  |
| 2019 | 18,339 (62.6) | 7,579 (25.9) | 3,367 (11.5) |
| 2020 | 21,185 (72.0) | 5,911 (20.1) | 2,324 (7.9) |
| 2021 | 22,218 (74.8) | 5,624 (18.9) | 1,852 (6.3) |
| 2022 | 13,372 (76.8) | 3,008 (17.3) | 1,032 (5.9) |
| Age, mean (SD) | 28.3 (5.68) | 28.1 (5.75) | 27.8 (5.77) |
| Residency, n (%) |  |  |  |
| Urban | 55,531 (73.9) | 16,495 (74.6) | 6,325 (73.8) |
| Rural | 19,578 (26.1) | 5,625 (25.4) | 2,250 (26.2) |
| Unknown | 5 (0.01) | 2 (0.01) | 0 (0.0) |
| Education, n (%) |  |  |  |
| Primary | 4,470 (6.0) | 1,501 (6.8) | 731 (8.5) |
| Secondary | 30,879 (41.1) | 9,594 (43.4) | 3,687 (43.0) |
| Higher | 27,221 (36.2) | 7,330 (33.1) | 2,479 (28.9) |
| Unknown | 12,521 (16.7) | 3,697 (16.7) | 1,678 (19.6) |
| ANC visits, n (%) |  |  |  |
| < 4 | 1,676 (2.2) | 556 (2.5) | 321 (3.7) |
| 4-8 | 55,742 (74.2) | 15,980 (72.2) | 6,037 (70.4) |
| >8 | 17,696 (23.6) | 5,586 (25.3) | 2,217 (25.9) |
| BMI, n (%)<sup>1</sup> |  |  |  |
| <18.5 | 4,987 (6.8) | 1,565 (7.3) | 610 (7.5) |
| 18.5-24.9 | 43,338 (59.4) | 12,948 (60.7) | 5,077 (62.1) |
| 25-30 | 16,052 (22.0) | 4,618 (21.7) | 1,646 (20.2) |
| >30 | 8,594 (11.8) | 2,194 (10.3) | 839 (10.3) |
| Parity, n (%) |  |  |  |
| Nullipara | 31,606 (42.1) | 8,130 (36.8) | 2,913 (34.0) |
| Multipara | 43,508 (57.9) | 13,992 (63.2) | 5,662 (66.0) |
| Plurality, n (%) |  |  |  |
| Singleton | 74,424 (99.1) | 21,887 (99.0) | 8,455 (98.6) |
| Multiple | 690 (0.9) | 235 (1.0) | 120 (1.4) |
| GA at lowest Hb value, mean (SD) | 35.5 (1.70) | 34.9 (2.27) | 34.7 (2.47) |
| Lowest recorded Hb value, mean (SD) | 118 (0.83) | 104 (0.31) | 90 (1.00) |
| Bleeding during pregnancy, n (%) |  |  |  |
| Yes | 1,166 (1.6) | 451 (2.0) | 165 (1.9) |
| No | 73,948 (98.4) | 21,671 (98.0) | 8,410 (98.1) |
| Mode of delivery, n (%) |  |  |  |
| CS | 30,202 (40.2) | 9,031 (40.8) | 3,656 (42.6) |
| Vaginal | 44,912 (59.8) | 13,091 (59.2) | 4,919 (57.4) |

<sup>1</sup> 3,343 observations are not having BMI measures

**Table 2.** Unadjusted and aORs and 95% CIs for the association between anemia during pregnancy and post-delivery ICU admission and preterm delivery.

|  | Non anemia<br>(reference<br>group)<br>n=75,114 |  | Mild 100-109 g/L<br>n=22,122 |  |  | Moderate & Severe <99 g/L<br>n=8,575 |  |  | p-value for<br>linear trend |
| --- | --- | --- | --- | --- | --- | --- | --- | --- | --- |
| Outcome | n (%) | OR/aOR | n (%) | OR<br>(95%CI) | aOR (95%CI) | n (%) | OR<br>(95%CI) | aOR (95%CI) |  |
| Post-delivery<br>ICU<br>admission | 377<br>(0.5) | 1.0/1.0 | 129<br>(0.6) | 1.16<br>(0.95-1.42) | 1.19 (0.97-1.46) | 52 (0.6) | 1.21 (0.90-1.62) | 1.24 (0.93-1.66) | 0.13 |
| Preterm<br>delivery | 2,436<br>(3.2) | 1.0/1.0 | 699<br>(3.2) | 0.97<br>(0.89-1.06) | 0.95 (0.87-1.04) | 276<br>(3.2) | 0.99 (0.87-1.13) | 0.93 (0.82-1.06) | 0.01 |

Among women with at least one Hb measurement in the third trimester, 558 were admitted to the post-delivery ICU, and the proportions were relatively similar across the anemia groups (no anemia:0.5%, mild and moderate/severe anemia:0.6%) (Table 3). Furthermore, 3,411 women delivered preterm, and the proportion of preterm deliveries was similar in all exposure groups (3.2% in the non-anemia, mild, and moderate/severe anemia groups). After adjustments for confounding factors, women with mild or moderate/severe anemia experienced higher odds of post-delivery ICU admission than the reference group (aOR, 1.19; 95% CI, 0.97–1.46 and aOR, 1.24; 95% CI, 0.93– 1.66, respectively), although the association was non-significant with a .05 threshold, and there was no sufficient evidence for a linear trend (*P* for trend = .13). In contrast, we observed a linear inverse association between anemia and preterm delivery (mild: aOR, 0.95; 95% CI 0.87-1.04; moderate and severe: aOR, 0.93; 95% CI 0.82-1.06, respectively, and *P* for trend= .01).

## Discussion

This is the first national birth registry-based study to describe the prevalence of anemia in pregnant women in Georgia and evaluate its association with selected adverse maternal outcomes. According to Hb measurements in the GBR, 40.6% of pregnant women in Georgia who delivered between January 1, 2019, and August 31, 2022, had anemia at least once during their pregnancy. Adjara in Western Georgia had the highest prevalence of women with anemia (47.0%), whereas Samtskhe-Javakheti in the south had the lowest prevalence (25.1%). Our results suggest that the overall prevalence of anemia during pregnancy is slightly higher than the global estimate of anemia in pregnant women (36.5%) in 2019 provided by World Bank Health Nutrition and Population Statistics [33]. Moreover, the same source indicated that the prevalence in Georgia is higher than those of neighboring countries, Armenia (18.1%) and Azerbaijan (35%). According to the 2019 WHO Global Health Observatory Data Repository, the prevalence of anemia in pregnant women in Georgia is higher than the average prevalence for upper-middle-income countries in Europe (24.5%) and substantially higher than those of high-income countries (17.2%) [34]. There is a considerable difference in the prevalence of anemia between the regions of Georgia. Accordingly, the presented prevalence for Adjara (47.0%), Kvemo Kartli (46.5%), Imereti (44.1%) and Guria (42.6%) are close to the anemia prevalence in low-income countries (42.6%) whereas the anemia prevalence in Samtskhe-Javakheti (25.1%) is closer to the prevalence in WHO European region (23.5%) [33, 34]. Specific dietary practices and differences in nutrition are possible explanations for the observed regional differences. For instance, in Western Georgia, red meat is less commonly consumed compared to the other parts of the country; this can partly explain the substantially higher prevalence of anemia in Adjara and Imereti than in Samtskhe-Javakheti.

The proportion of women with mild (28.3%), moderate (11.9%), and severe (0.5%) anemia in Georgia was consistent with pooled estimates from Canada and China [8, 9, 12, 18], and lower than that the reported values for some African and Asian countries, such as Somalia, India, and Pakistan [10, 35]. The low prevalence of severe anemia suggests that implemented national public health measures for anemia prevention, such as free screening and treatment among pregnant women, are efficient. Women with anemia were also more likely to have more than eight ANC visits than women without anemia, indicating that women who have been screened and diagnosed with anemia receive more intense follow-up and treatment than others, which might have resulted in the low observed prevalence of severe anemia.

Another important finding of this study is that 18.1% of all pregnant women who visited the ANC had no reliable Hb measurement during their entire pregnancy, and this proportion varied considerably across the country (Supplementary Table2). This may impact statistics on the true prevalence of anemia in the country and in certain regions. According to the State Antenatal Care Guidelines, all pregnant women attending ANC should have at least one Hb measurement [27]. Hence, the above proportion of women without Hb measurements indicates that despite state-supported ANC and free access to medication for the treatment of iron deficiency anemia during pregnancy (if detected before GA week 13), almost one-fifth of all pregnant women may not receive the care they require or if they do, it may not be registered in the GBR, although this registration is mandatory for ANC providers. These findings suggest that a major priority for Georgian decisionmakers should be to identify the reasons behind the lack of Hb measurements, resolve problems with the registration of implausible values, and follow up women with no Hb measurements to ensure equal access to high-quality ANC for all pregnant women across the country.

Consistent with previous reports, this registry-based study demonstrated an association between anemia during the third trimester and increased odds of admission to the post-delivery ICU [9, 35], although this was not statistically significant using a 5% threshold (*P* for trend = .13). Anemia in pregnancy is a well-known risk factor for post-partum hemorrhage [11], which leads to post-delivery ICU admission. Our results should be interpreted with caution but are notable since hemorrhage has been identified as the leading direct cause of post-delivery ICU admission in Georgia, eventually leading to maternal death [25]. Timely and properly initiated preventive interventions are required during ANC to reduce the risk of maternal morbidity and mortality in Georgia, especially since the maternal mortality rate has increased since 2020 [23].

In contrast to previous studies [3, 8, 10, 35, 36], we found no increased odds for preterm delivery among mothers with anemia in the third trimester, and this is a good message for decisionmakers of the Georgian national healthcare. In fact, we observed an inverse linear trend, suggesting that women with anemia have lower odds of preterm delivery than women without anemia (*P* for trend =0.01). This seems contradictory because several other studies have reported an increased risk of preterm delivery [2, 4]. However, the risk seems to differ according to trimester and across studies; for example, a systematic review and meta-analysis from South Africa, published in 2022 highlights difficulties in clarifying the association between anemia and preterm delivery and in which direction the association might be [12]. Similarly, meta-analyses published in 2000 [37] and 2019 [7] reported a non-significant inverse relationship between anemia in late pregnancy and preterm delivery, which is in accordance with our results. One possible explanation for this difference could be that women in middle-income countries with a well-developed registration system receive intensive follow-up and treatment if they are diagnosed with anemia, while women in lower-income countries with limited capacities in the healthcare sector may not receive optimal follow-up and medical care due to a lack of resources or different screening approaches [8]. Our results show a higher frequency of ANC in pregnant women with a low Hb value. Thus, with special care and treatment, anemia-related preterm delivery is an avoidable adverse maternal outcome, and anemia could be effectively managed even during the third trimester of pregnancy.

The substantial differences in the regional prevalence of anemia in Georgia highlight the need for a customized and stronger public health response and policy implementation toward anemia as a significant public health challenge in Georgia. The proportions of pregnant women who do not attend ANC and of those with no registered Hb values indicate the need for a coordinated strategy to ensure that women in Georgia have equal access and receive optimal care independent of their place of residence. Moreover, it is advisable to evaluate the registration structure of service-related information in the GBR. Nevertheless, focusing on increased, countrywide coverage of the state ANC program with all its features, better access to optimal ANC, and coordinated customized, multilevel responses against anemia are important steps toward mitigating the consequences of maternal anemia and equalizing regional disparities in anemia prevalence.

### Strength and limitations

The strength of this study is the large number of women we included, which makes the study representative of pregnant women in Georgia. However, almost one-fifth of the women did not have a valid Hb measurement. Women who do not attend ANC may have a higher risk of anemia because they do not receive proper follow-up from medical personnel. The significant number of pregnant women not attending ANC may have affected the prevalence estimate in the present study. Consequently, there is a need to improve ANC operations to obtain reliable estimates of the prevalence of anemia in pregnant women in Georgia. Based on the GBR data, most women underwent more than one Hb measurement. However, we classified the women according to the lowest registered Hb value and did not consider whether the women were treated or whether the Hb level of those who were treated normalized. This is a limitation of the study. In addition, anemia was identified using measured Hb values, and we could not specify the type of anemia. Moreover, despite controlling for many covariates in the multivariate-adjusted regression analyses, measurement errors could have resulted in residual confounding, and we had no information about several important factors, especially comorbidities such as chronic kidney disease, infections, and cancer, which could have influenced the results.

Despite these limitations, this study provides important insight into the prevalence and potential consequences of anemia in pregnant women in Georgia.

## Conclusion

Approximately one-third of pregnant women in Georgia experience anemia at any time during pregnancy, but severe anemia is rare. One-fifth of the women who attended ANC did not have any valid Hb measurement, which is surprising and worrisome since Hb measurements are covered by the state of Georgia. There were large regional differences in the prevalence of anemia, which warrant further investigation. To accelerate progress toward sustainable development goals and decrease the public health burden of anemia in Georgia, early identification and adequate management of anemia during pregnancy are crucial.

## Acknowledgments

The authors would like to acknowledge the assistance of the Institute of Hematology and Transfuziology in providing important insights regarding the study design.

Moreover, the authors would like to thank Editage (www.editage.com) for English language editing.

The publication charges for this article have been funded by a grant from the publication fund of UiT The Arctic University of Norway.

## Author contribution

All authors contributed to the methodology, analysis, writing, and editing of the manuscript. All authors approved the final version of the manuscript and agreed to be responsible for all provided information. CR, TM, TB, EA and AG supervised the study. NS, CR, and TM summarized the national datasets and contributed to data collection and curation. NS drafted the manuscript and amended it according to feedback from all authors.

## Consent for publication

Not applicable.

## Availability of data and materials

According to Georgian legislation, data from central health registries can be used for research purposes as long as personal data is not shared. Data collected for this study are not publicly available because they contain personalized and potentially identifying information. Proper ethical and legal approvals are required for access before requesting the data, and only anonymous/pseudonymized data can be shared. Researchers can contact the National Center for Disease Control and Public Health in Georgia or the corresponding author for assistance on how to apply for access to data from registry.

## Supporting information

**Figure 1.**

**Figure 2.**

**S1 File.**

**S2 File.**

**S1 Table.**

**S2 Table.**

**S3 Table.**

